# Corr-A-Net: Interpretable Attention-Based Correlated Feature Learning framework for predicting of HER2 Score in Breast Cancer from H&E Images

**DOI:** 10.1101/2025.04.22.25326227

**Authors:** Kaushik Dutta, Debojyoti Pal, Suya Li, Chandresh Shyam, Kooresh I. Shoghi

## Abstract

Human epidermal growth factor receptor 2 (HER2) expression is a critical biomarker for assessing breast cancer (BC) severity and guiding targeted anti-HER2 therapies. The standard method for measuring HER2 expression is manual assessment of IHC slides by pathologists, which is both time intensive and prone to inter- and intra-observer variability. To address these challenges, we developed an interpretable deep-learning pipeline with Correlational Attention Neural Network (Corr-A-Net) to predict HER2 score from H&E images. Each prediction was accompanied with a confidence score generated by the surrogate confidence score estimation network trained using incentivized mechanism. The shared correlated representations generated using the attention mechanism of Corr-A-Net achieved the best predictive accuracy of 0.93 and AUC-ROC of 0.98. Additionally, correlated representations demonstrated the highest mean effective confidence (MEC) score of 0.85 indicating robust confidence level estimation for prediction. The Corr-A-Net can have profound implications in facilitating prediction of HER2 status from H&E images.

## INTRODUCTION

Breast cancer (BC) is regarded as one of the most widespread cancers worldwide, contributing to the second leading cause of cancer-related mortality in women (1, 2). BC exhibits significant heterogeneity, which underscores the need for conducting pathological assessment for tumor histology to determine optimal treatment planning (3, 4). The human epidermal growth receptor 2 (HER2) oncogene, also known as ERBB2 or neu, encodes the type I receptor tyrosine kinase HER-2/neu promotes cell growth and is typically found in cell membranes. Approximately 15-20% of BC patients demonstrate HER2 gene amplification or overexpression, which is associated with tumor aggressiveness, proliferation and resistance to conventional therapies (5-8). The development of targeted anti-HER2 therapies (acting as HER2 inhibitors), such as using trastuzumab (9), lapatinib (10) and petuzumab (11) has shown to decrease patient mortality and increase outcome (12, 13). In order to administer these treatment strategies accurate measurement of HER2 expression in breast cancer is crucial.

In clinical practice, the HER2 score is assessed by performing Immunohistochemical (IHC) staining, followed by pathologists analyzing tissue samples manually to determine the protein expression level. Hematoxylin and eosin (H&E) counterstaining are employed for the cell nuclei and other tissue features. Machine learning coupled with digital slide scanners have enabled automated analysis of Whole Slide Images (WSIs) for predicting disease characteristics by analyzing complex cellular and protein characteristics and widely applied in breast cancer (14-19). Deep learning methods have the capability to capture multi-scale intricate details from WSIs and find applications in cell segmentation (20), tumor classification (21), biomarker estimation (22, 23) and cancer localization (24). In HER2 score prediction, deep-learning methods are employed at both the slide level and the patch level on immunohistochemistry (IHC) stained slides, enabling a comprehensive analysis of HER2 expression and enhancing predictive accuracy (25-29). H&E slides are also used extensively to predict HER2 score leveraging deep-learning pipelines (30-34). Despite these advancements, the absence of biologically interpretable morphological features in H&E-stained slides limits their utility for the quantitative prediction of HER2 expression levels (35). In recent times, the development of Correlational Neural Network (CorrNet) (36) has shown capabilities to produce common representations from multi-modal data by leveraging concept of canonical correlation and multi-modal autoencoder from unimodal data post training.

In this work we developed a two-step deep-learning based framework to predict the HER2 level expression from H&E slides. The first step employs a Correlational Attention Network (Corr-A-Net) that leverages spatially aligned H&E and IHC slides from breast cancer patients to learn the correlated features between these two imaging modalities. The Corr-A-Net exhibits multi-modal image features fusion using an attention mechanism. In the second step, a predictive network is trained using the correlated representations derived exclusively from H&E images to forecast HER2 expression levels. For the predictive network, we implemented Vision Transformer (37), EfficientNet (38), and ResNeXT-50 (39) architectures, alongside a confidence score predictor that assigns a learnable confidence score to each prediction. Additionally, we leveraged the potential of explainable artificial intelligence by applying Gradient Class Activation Maps (Grad-CAM) (40) to identify the morphological features contributing to the predictive decisions. The overview of the framework is given in Figure. 1.

**Figure 1.**
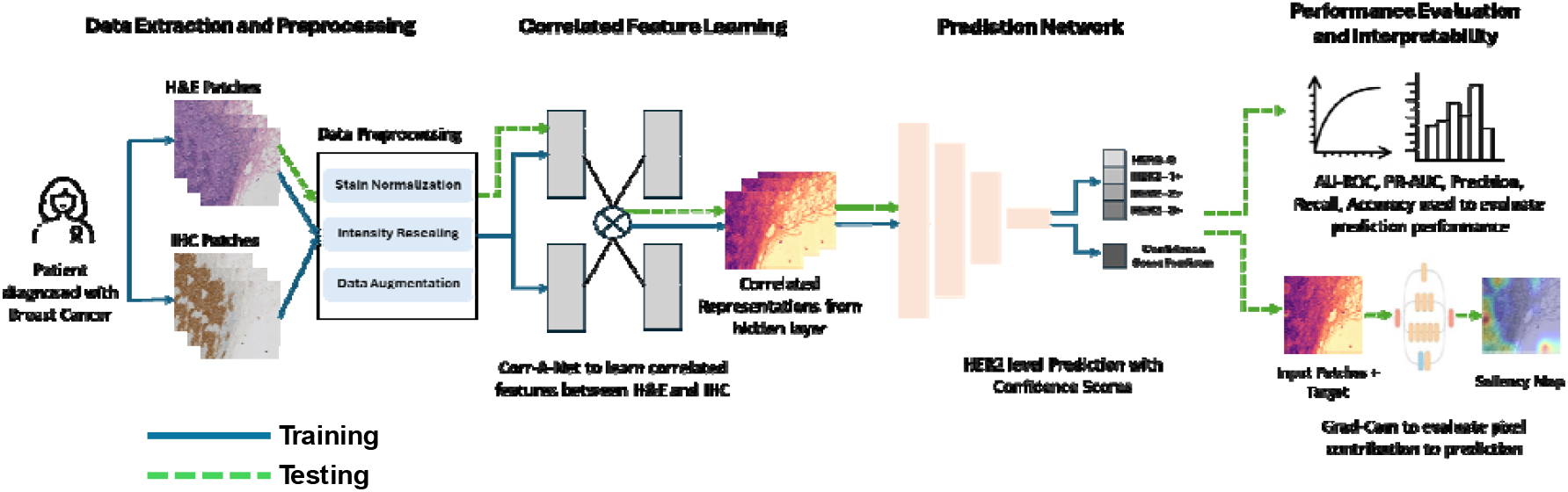
Overview of the proposed framework to predict HER2 score using correlated feature learning

## RESULTS

### Degree of Correlation for Latent Representations

Figure 2A depicts the representative patches from IHC, H&E and hidden/latent representations for different HER2 level expression generated by the Corr-A-Net model. These correlated representations were generated using post-trained Corr-A-Net model from H&E images only. The correlated representations visibly capture important imaging features, particularly the membranous structures characteristic in IHC images which are indicator of HER2 level prediction. This highlights Corr-A-Net’s ability to effectively translate imaging features between modalities during training which can be leveraged during inferencing using unimodal data along with giving a dimension of interpretability during prediction.

**Figure 2.**
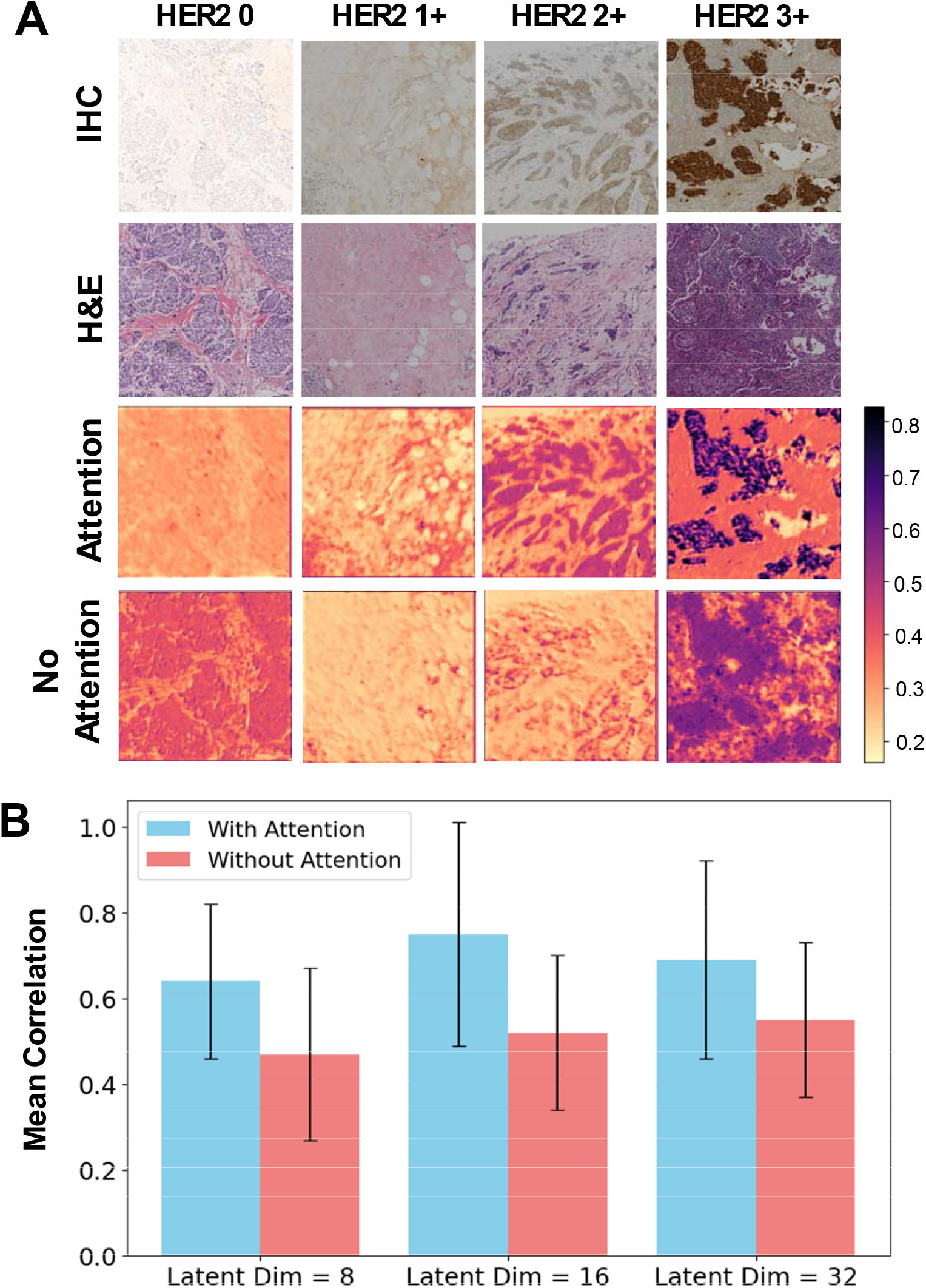
**(A)** Correlated Representations **(B)** Mean Correlation between the hidden representations and the IHC images for different filter sizes (8, 16, 32) of Corr-A-Net for both with and without attention scenarios

The ability of Corr-A-Net to learn correlated representations from unimodal data (i.e. H&E data only) is evaluated by computing the mean correlation between the hidden representations and the IHC images. We evaluated the mean correlation performance for two different scenarios of Corr-A-Net i.e. with and without using the squeeze-excitation attention block and is depicted in Figure 2B. For three different latent dimensions i.e. 8, 16 and 32 we observed that the mean correlation is maximum for dimension = 16 both attention (ρ = 0.76) and dimension = 32 for without attention (ρ = 0.55) scenario. The shared representations using the SA block (i.e. using attention mechanism) in the Corr-A-Net exhibited significantly better (*p*≤*0*.*05, Wilcoxon Test*) mean correlation score compared to without using the SA block.

### HER2 level Prediction Performance

The HER2 level prediction performance using of the deep-learning based classifiers (ViT, EfficientNet, ResNeXT) were evaluated using BA, WP, WR and MCC on the independent testing dataset (978 patches). The performance of the correlated representations obtained from H&E only in predicting HER2 level was benchmarked against classification using H&E slides alone, IHC slides alone, as well as combined IHC and H&E slide inputs, providing a comparative analysis of modality-specific and multimodal prediction approaches. The prediction performance of the classifiers is summarized in Table. 1.

**Table. 1.** HER2 Expression Level Prediction Performance results using WP, WR, BA, MCC, AUC and PR-AUC for different models.

|  | Model | WP | WR | BA | MCC | AUC | PR-AUC |
| --- | --- | --- | --- | --- | --- | --- | --- |
| H&E Only | EfficientNet | 0.69 | 0.64 | 0.60 | 0.64 | 0.77 | 0.70 |
|  | ViT | 0.72 | 0.75 | 0.72 | 0.65 | 0.82 | 0.75 |
|  | ResNeXT | 0.79 | 0.77 | 0.80 | 0.77 | 0.91 | 0.84 |
| IHC Only | EfficientNet | 0.74 | 0.73 | 0.63 | 0.66 | 0.83 | 0.76 |
|  | ViT | 0.82 | 0.80 | 0.76 | 0.71 | 0.88 | 0.80 |
|  | ResNeXT | 0.87 | 0.88 | 0.85 | 0.81 | 0.94 | 0.91 |
| H&E and IHC Combined | EfficientNet | 0.87 | 0.86 | 0.87 | 0.80 | 0.84 | 0.73 |
|  | ViT | 0.88 | 0.87 | 0.88 | 0.82 | 0.94 | 0.88 |
|  | ResNeXT | 0.91 | 0.90 | 0.92 | 0.86 | 0.98 | 0.96 |
| Corr-A-Net (Filter Size = 8) (H&E Only) | EfficientNet | 0.86 | 0.85 | 0.84 | 0.78 | 0.84 | 0.72 |
|  | ViT | 0.86 | 0.86 | 0.87 | 0.79 | 0.95 | 0.89 |
|  | ResNeXT | 0.88 | 0.88 | 0.89 | 0.83 | 0.97 | 0.94 |
| Corr-A-Net (Filter Size = 16) (H&E Only) | EfficientNet | 0.88 | 0.87 | 0.83 | 0.82 | 0.88 | 0.76 |
|  | ViT | 0.86 | 0.85 | 0.89 | 0.78 | 0.95 | 0.9 |
|  | ResNeXT | 0.91 | 0.90 | 0.94 | 0.87 | 0.98 | 0.95 |
| Corr-A-Net (Filter Size = 32) (H&E Only) | EfficientNet | 0.87 | 0.87 | 0.84 | 0.81 | 0.88 | 0.75 |
|  | ViT | 0.86 | 0.87 | 0.89 | 0.81 | 0.94 | 0.88 |
|  | ResNeXT | 0.93 | 0.92 | 0.93 | 0.90 | 0.97 | 0.96 |

The HER2 level prediction using H&E alone exhibited the lowest performance across all evaluated network models, achieving a BA score of 0.60, 0.72 and 0.80 for EfficientNet, ViT and ResNeXT respectively. The IHC alone achieved a BA score of 0.63, 0.76, and 0.85 for EfficientNet, ViT and ResNeXT respectively. However, when the IHC patches were integrated with H&E patches, the performance improved significantly, and the BA increased by 0.24, 0.12, and 0.07 for EfficientNet, ViT, and ResNeXT, respectively, relative to the IHC-only approach. These results demonstrate that multimodal integration of H&E and IHC patches significantly improves classification performance compared to unimodal data. The correlated representations generated by Corr-A-Net by utilizing H&E patches alone were assessed for predicting HER2 level expression across three distinct latent dimensions i.e. 8,16 and 32. In all cases, the representations exhibited significant improvement in HER2 level prediction compared to the IHC only model. Furthermore, when compared to the performance of models leveraging combined H&E and IHC patches, the correlated representations yielded a marginally better BA score for ResNeXT and ViT models using latent dimensions of 16 and 32. Overall, the ResNeXT consistently demonstrated superior BA scores compared to other classifier models across all evaluated latent dimensions.

Figure. 3 depicts the Receiver Operating Characteristics (ROC) curve and the AUC values for IHC only, IHC and H&E combined and correlated representations in predicting HER2 level. The class-wise ROC was plotted to determine class-wise discriminative capability of the models for different scenarios. From Figure. 5 it is evident that for all network model the IHC and H&E combined, and the correlated representations AUC performance are better for all different classes compared to the IHC only model.

**Figure 3.**
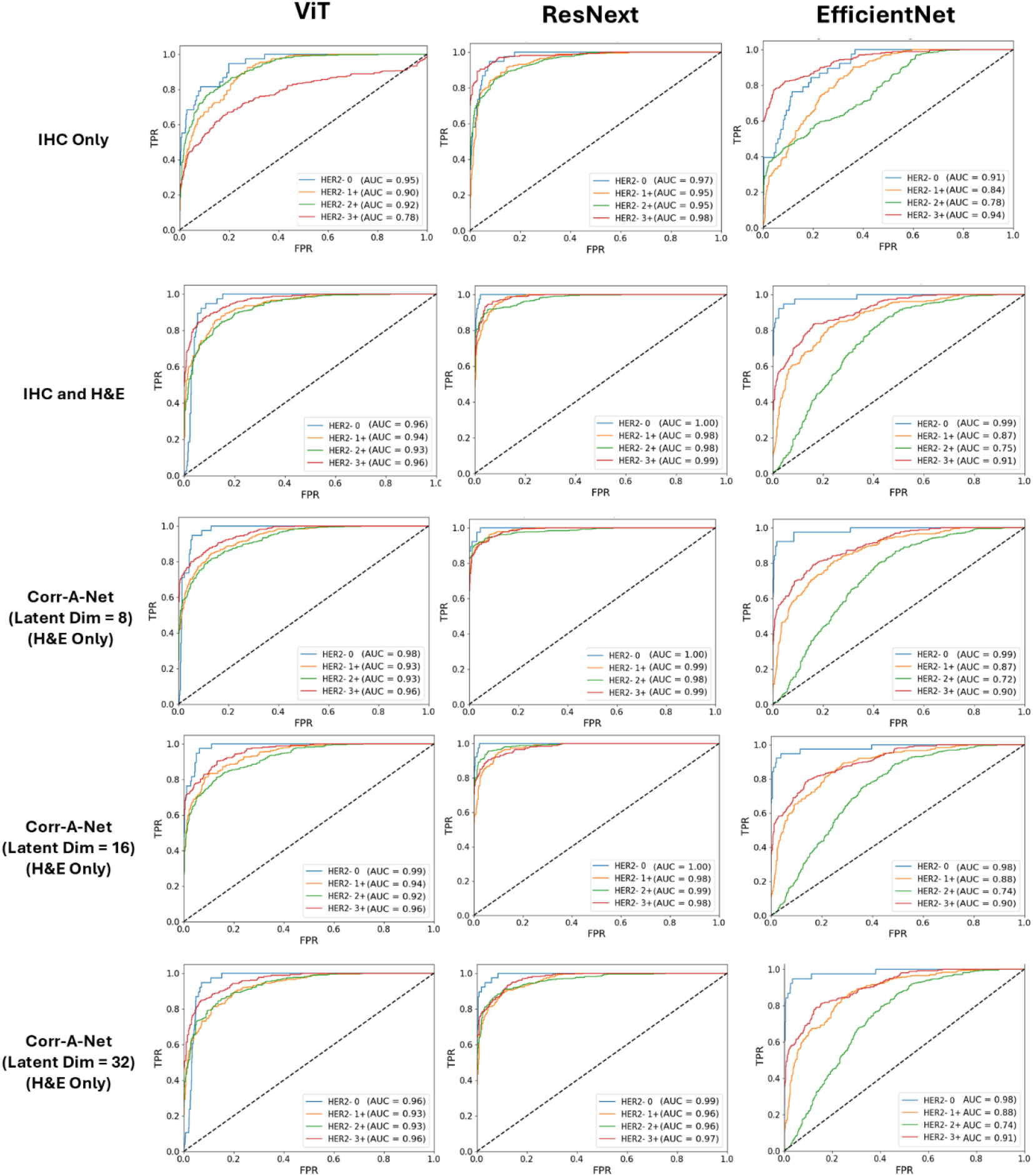
ROC curve and AUC values for EfficientNet, ResNeXT and ViT stratified for different HER2-level expression for IHC only, IHC and H&E combined and correlated representations.

### Confidence Score Evaluation

The accuracy of estimating confidence scores associated with HER2 prediction by the models is evaluated using the MEC metric. Figure 4A illustrates that predictions derived from correlated representations and those based on combined IHC-H&E inputs exhibited significantly improved MEC scores compared to predictions derived solely from IHC data. Particularly, ResNeXT based classification using correlated representations for latent dimensions of 16 and 32 exhibited the best MEC scores of 0.84 and 0.85 respectively. Consistent with the HER2 level prediction performance ResNeXT showed better MEC scores compared to its peer network models for all different settings.

**Figure 4.**
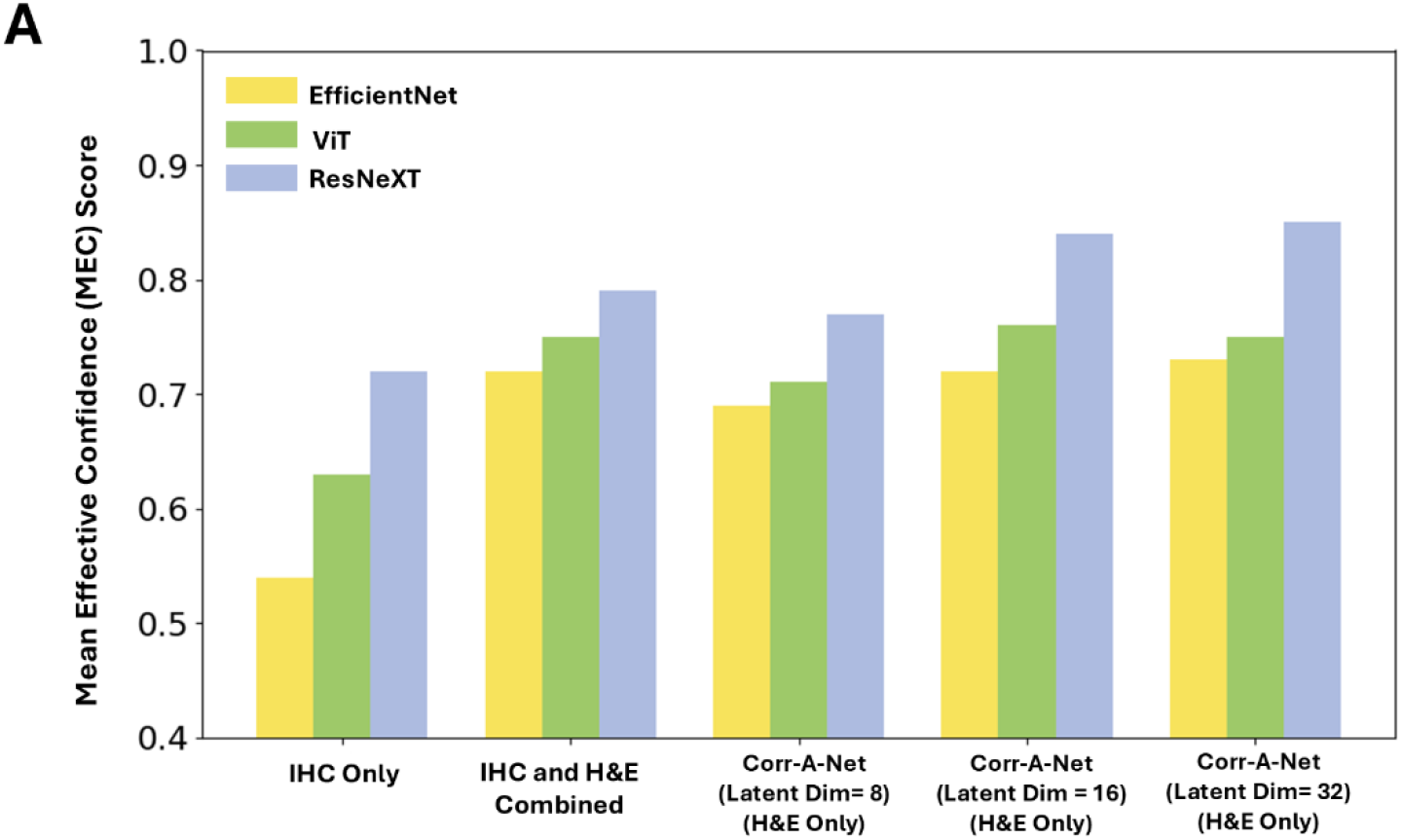
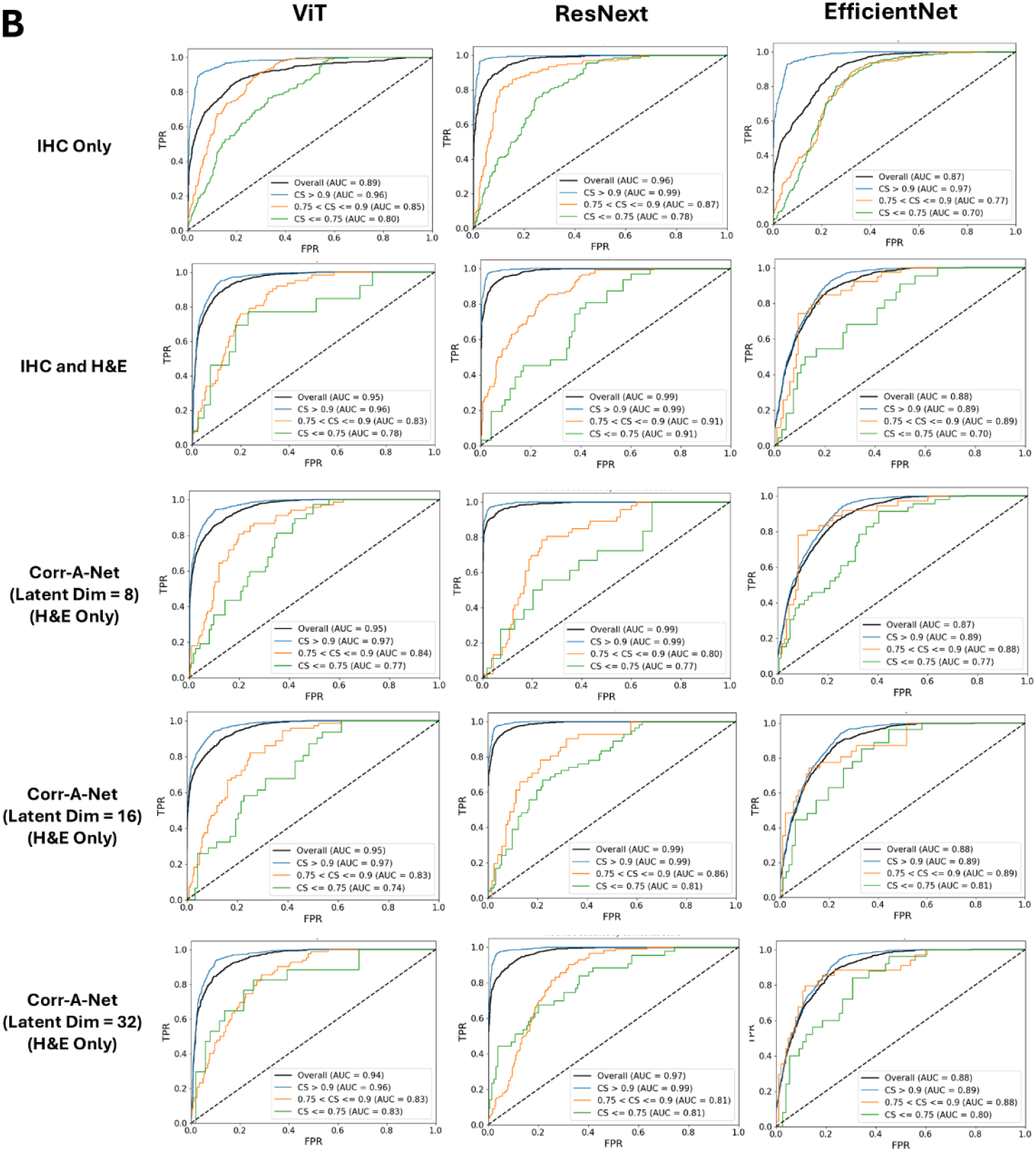
**(A)** Bar graph depicting the MEC score **(B)** ROC curve and AUC values for EfficientNet, ResNeXT and ViT stratified by different Confidence Score (CS)

Generally, higher MEC indicates that a greater proportion of correct predictions were made with high confidence score, while incorrect predictions were made with lower confidence score. The relationship of classification performance to the estimated confidence scores were evaluated by performing a stratification of patches based on the confidence score. Three level of stratification level were chosen i.e. for Confidence Score (CS) ≥ 0.9 (high confidence), 0.75 ≤ CS < 0.9 (medium confidence), CS < 0.75 (low confidence) and the classification performance was evaluated for all stratification level using ROC analysis (Figure. 4B). The AUC score was consistently higher for all the scenarios when the CS was greater than 0.9 suggesting that the confidence predictor network consistently predicts correct confidence level for correct predictions.

### Explainability Analysis

Figure 5A depicts the Grad-CAM map to assess interpretability of the prediction network in two different scenarios i.e. when using correlated representations subjected to attention block and when not subjected to attention block. The visual inspection of the Grad-CAM maps suggests that attention based correlated representations have better ability of localizing the relevant areas for prediction when compared to the no-attention based correlated representations. The Grad-CAM’s performance in generating relevant explanation maps were evaluated using the Drop in Accuracy metric as shown in Figure 5B. The predictions derived from attention based correlated representations had consistently lower percent of Drop in Accuracy for all the latent dimensions compared to the no-attention scenario. The most effective explanation maps were obtained utilizing ResNeXT model with latent dimension of 16, yielding a drop in accuracy of only 20.32%, while the ViT model for the same configuration achieved slightly higher drop of 22.24%. This suggests that attention mechanism can effectively guide the model to focus on regions which have higher likelihood of contributing to accurate predictions.

**Figure 5.**
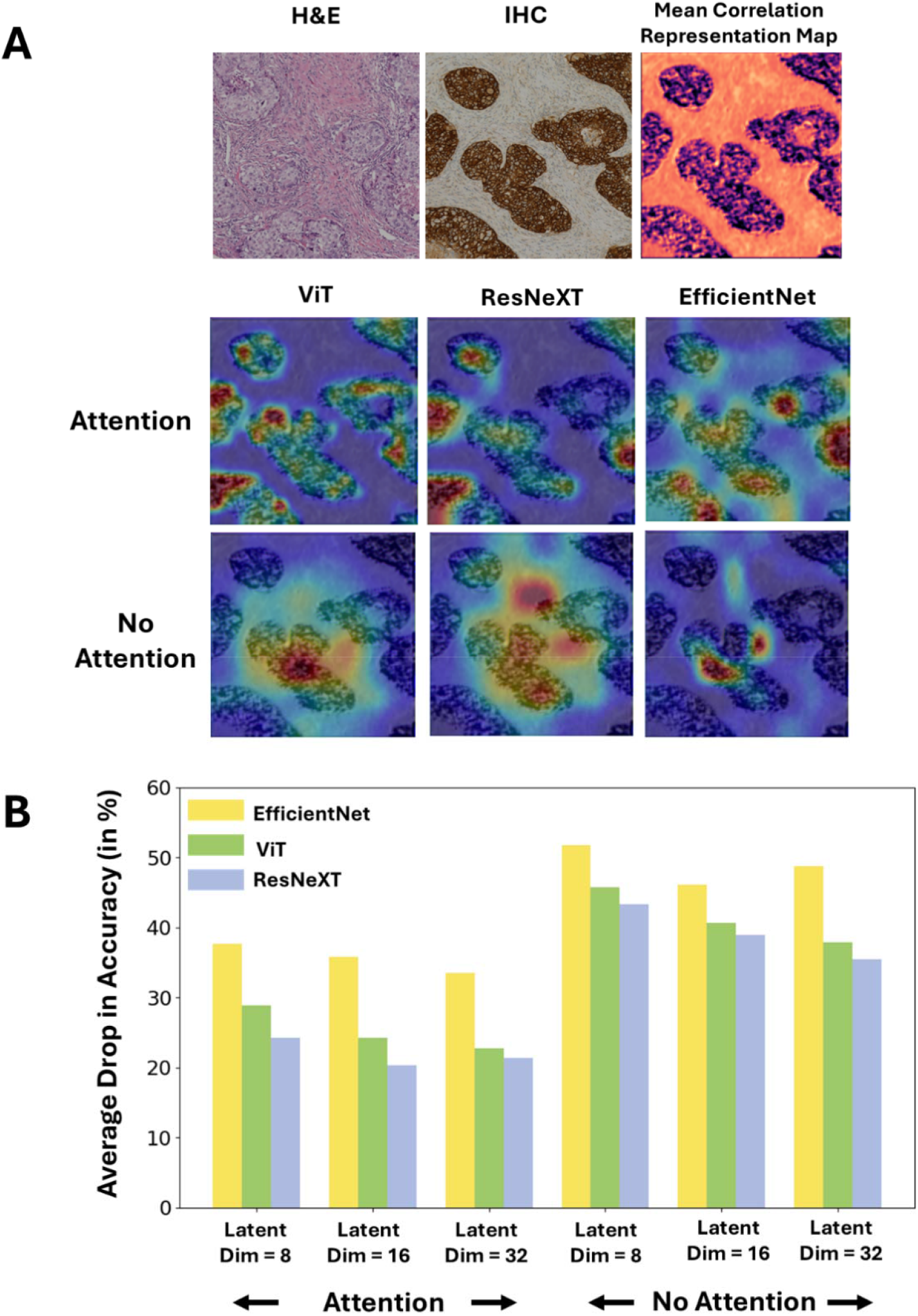
**(A)** Explainability maps obtained using Grad-CAM for ViT, ResNeXT and EfficientNet using attention based and no-attention based correlated representation **(B)** Bar-Graph depicting the Drop in Accuracy (in %) for different prediction network under attention and no-attention scenario.

## DISCUSSION

The objective of this work was to develop a framework to predict the HER2 expression score by utilizing only H&E images. To that end, we developed a novel Corr-A-Net architecture which is trained using IHC and H&E images to generate correlated representations. Though Corr-A-Net was trained jointly on both modalities, it is capable of generating correlated representations from H&E images alone in post-training phase. Importantly, the correlated representations generated by Corr-A-Net incorporate IHC-based features, thereby providing biological significance to the framework, as demonstrated by our interpretability analysis. The H&E derived correlated representations are subsequently leveraged by three distinct classifier networks to predict HER2 level expression. Additionally, our developed framework includes a confidence prediction module that aids in stratification of cases and enhances interpretability. Previous state-of-the-art approaches for HER2 prediction from H&E images included HAHNet (32), which employed a parallel InceptionV3 architecture with channel attention blocks, and HE-HER2Net (41), a transfer learning-based model utilizing XceptionNet. In comparison, our pipeline achieved the highest overall ROC-AUC score of 0.98 and a balanced accuracy of 94.26% for ResNeXT (latent-dim = 16) in predicting HER2 status from derived correlated representations, which is a 7.5% and 1.5% improvement over HE-HER2Net and HAHNet performance. Moreover, these networks did not incorporate the confidence predictor component or the interpretability analysis to signify biological relevance.

Clinically, HER2 status is assessed using IHC images; however deep-learning based methods have utilized either IHC or H&E separately to predict HER2 scores (15, 16, 21, 25, 26, 28, 30-35). However, H&E images exhibit distinct morphological features compared to IHC and do not capture the HER2 protein expression which is essential for clinical interpretation of HER2 level prediction. For this reason, we trained the Corr-A-Net which effectively learns the correlated feature space between both modalities such that post-training it can be leveraged to generate correlated representations from unimodal data (H&E only). Recent efforts have established that common representation learning through correlational neural networks is capable of generating correlated representation between histopathology (H&E) and radiology (MRI) using MRI data only, facilitating cancer detection/segmentation and interpretability (42, 43). The integration of the attention mechanism in the Corr-A-Net enables the model to compute weighted feature maps by selectively emphasizing critical spatial features while supressing less informative ones. The attention mechanism included within CNN model has demonstrated to produce better classification results for natural (44) and medical images (45, 46) which was persistent in our study as the correlated representations from attention based Corr-A-Net outperformed the no-attention based representations in terms of predictive performance.

Most HER2 prediction algorithm evaluation rely on traditional classification metrics, undermining the quantitative reliability of the prediction. Confidence scores, which quantifies the probability of predictive correctness have been extensively used object detection/segmentation though frameworks like R-CNN and YOLO (47-49). In image classification, the distribution and entropy of the class probabilities are utilized to derive confidence estimates (50, 51). Recent advancements in pathology image analysis have applied confidence estimation through using Multiple Instance Learning (MIL) (52) and ensemble models (53). In MIL, the confidence scores are computed from the probability of an instance belonging to a specific bag of classes, while in ensemble models, confidence is calculated as the mean over precision scores for varying thresholds applied across multiple models. Borquez et al. implemented a Bayesian deep-learning classifier for predicting HER2 expression with Monte-Carlo dropout to simulate multiple classification outputs which are leveraged to model epistemic uncertainty and confidence in prediction (27). While these methods yield a confidence estimate, they are not learnable, meaning that the model predictions are not leveraged as feedback to further enhance performance. Wan et. al developed a confidence network (ConfNet) integrated within the CNN framework which automatically learns the confidence score in the training phase by utilizing negative correlation between model accuracy and task-loss (54). Building on this, we developed an incentivized mechanism for training our prediction models which incorporates a surrogate confidence network. The network learns to estimate confidence dynamically from prediction model’s performance, thus enabling learnable confidence. Our results demonstrated in Fig. 4 validates that low confidence is associated with lower AUC and lead to higher rate of misclassification, indicating that model exhibits more confidence for accurate predictions.

The inherent “black-box” nature coupled with limited interpretability and complexity of deep-learning based models limits their translation to clinical practices, as clinicians struggle to trust the reasoning of decision making (55, 56). Explainable artificial intelligence (XAI) which aims to provide task-based interpretation to model outcome has been popularized for medical image classification with the advent of class-activation maps (CAM) based techniques (57, 58). Interpretability is particularly critical in predicting HER2 level expression, as pathologists rely on assessing the intensity of protein expression within the membranous structure of IHC images. Previous work in HER2 level prediction have explored interpretability by analysing the spatial distribution of attention scores from MIL (59) and by computing the HER2 positive tumour cell percentage (60). Huang et. al have demonstrated that Grad-CAM ++ method revealed stronger influence in diseased tissue areas while predicting HER2 level from ultrasound images (61). These methods did not quantify how predictive performance changes with change in network model which is addressed in our framework by the utilization of drop in accuracy metric. Additionally, attention mechanism (by applying SA-block) has proven to further improve interpretability by guiding the model to focus on highly predictive markers during decision making (44). Our pipeline validated this effect in context of HER2 level prediction where the attention mechanism generated more informative interpretable maps with Grad-CAM than its non-attention counterpart (Figure 5).

Although the correlated representations generated from H&E showed promising and interpretable performance in predicting HER2 status, the framework has few limitations. First, the framework was trained and evaluated on a dataset obtained from the same slide scanner and from the same institution. It is well established that pathological patches vary significantly in terms of slide intensity and quality due to variability in acquisition protocols (62, 63). Although our algorithm incorporates intensity normalization and compute correlated representations in feature space to reduce slide specific variability. a comprehensive validation on external dataset containing pathological specimens acquired under different settings would enhance the generalizability of the network. Second, an ensemble-based approach rather than single network can further improve the predictive performance of the pipeline but at the cost of higher computation and training complexities. Finally, the explainability maps of the Grad-CAM method should be validated by manual pathologist delineation of predictive regions to make it more robust for clinical translation.

## METHODS

### Dataset and Preprocessing

We utilized the publicly available Breast Cancer Immunohistochemical (BCI) dataset (64) which consists of total 4873 spatially co-registered H&E and IHC image patches with a size of 1024 1024. The dataset was obtained using Hamamatsu NanoZommer S60 pathology section scanner at a scanning resolution of 0.46 μm per pixel from a total of 319 breast cancer patients. Each image patch was labelled for their respective HER2 level expression (0, 1+, 2+ and 3+). The interpretation of each HER2 level expression is given in Table. 2 and representative slide patches from the BCI datasets are depicted in Figure. 6. The dataset was split into training and testing with 3896 samples for training and 977 independent samples for testing the performance. The training samples were further split into training set and validation using 70:30 split for monitoring the performance of the network during training.

**Table 2.** HER2 Expression Level and their Interpretation.

| HER2 Expression Level/ IHC Score | HER2 Test Interpretation | HER2 Status |
| --- | --- | --- |
| 0 | No Staining or Incomplete and barely perceptible membrane staining in <10% tumor cells | Negative |
| 1+ | Faint and barely perceptible membrane staining in >10% of tumor cells; cells are reactive only in part of their membrane | Negative |
| 2+ | Weak to moderate complete basolateral or lateral membrane staining in $\geq 10\%$ of tumor cells | Equivocal |
| 3+ | Strong and intense complete basolateral or lateral membrane staining in $\geq 10\%$ of tumor cells | Positive |

**Figure 6.**
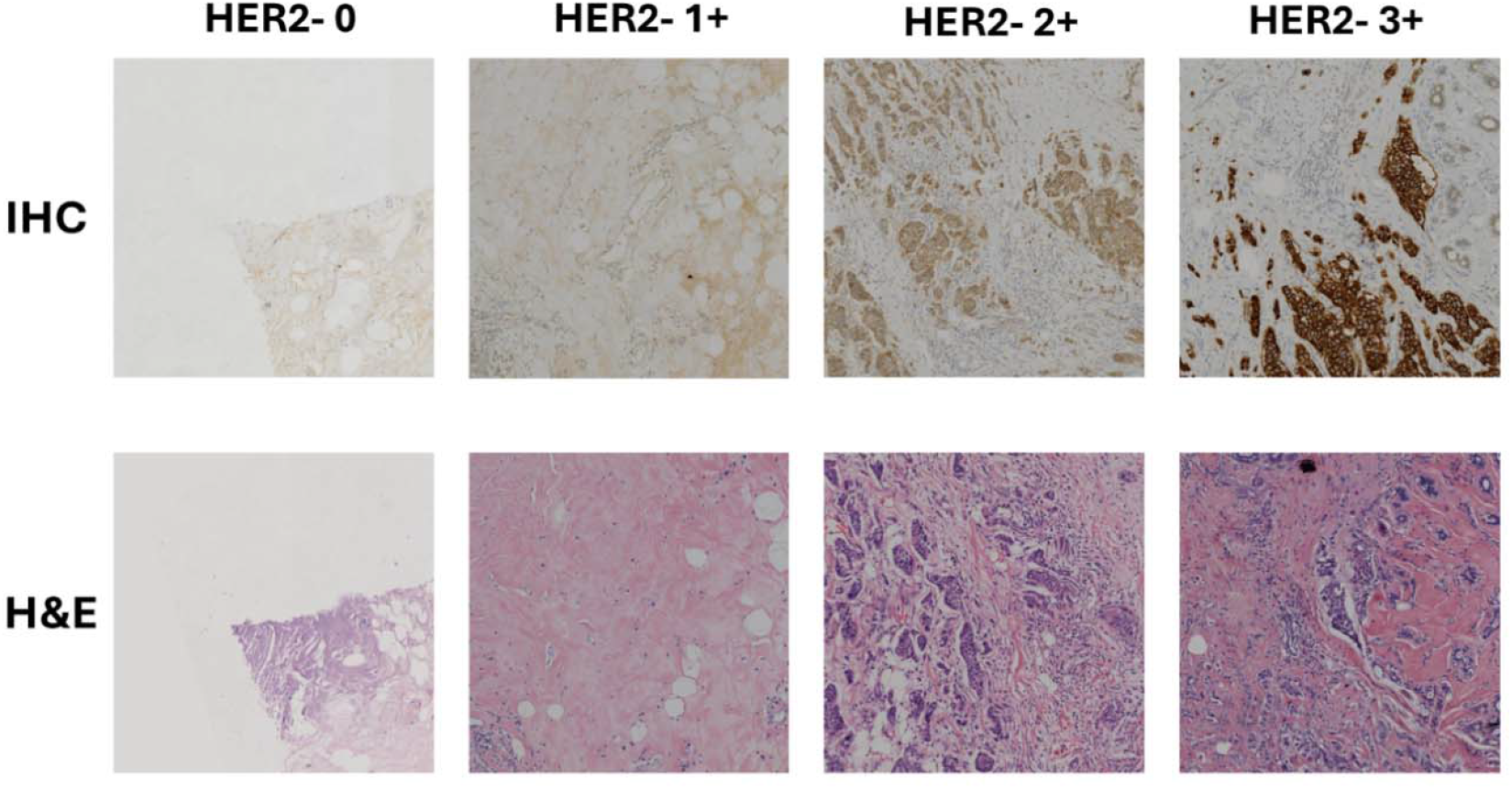
Representative slides from the BCI Dataset of H&E and IHC patches for different HER2 expression levels

For the data preprocessing step, the image patches were subjected to color normalization using Macenko Normalization to address the variability in staining processes across different tissue samples and slides (65). The image patches were resized to 256 256 pixels to reduce the computational processing times and to standardize input dimensions for the pretrained neural network architectures. The image patches intensity level was rescaled such that the intensity values are between 0 and 1. Data augmentation is applied in form of random rotation, intensity scaling, contrast adjustment and flipping to prevent overfitting of the model and increase model generalization.

### Correlational Neural Network

We developed a Deep Convolutional Correlational Attention Network (Corr-A-Net) model with three hidden layers having different filter sizes to learn the feature correlation between H&E and IHC patches. We further added a Squeeze-Excitation based attention mechanism in the innermost hidden layer to enhance the correlative feature learning.

#### Corr-A-Net Architecture

The deep convolutional Corr-A-Net is composed of three primary modules: feature extractor (encoder block), feature fusion (hidden layer with attention mechanism) and feature reconstructor (decoder block). The CorrNet is based on the principle of multi-modal autoencoders and canonical correlative analysis and aims to learn a common representation between two input vectors (here obtained from H&E and IHC) such that they are maximally correlated in latent space while at the same time minimizing the self and cross reconstruction loss. In the feature extraction step, the first convolutional block of the ResNet50 architecture (66) was utilized to extract feature maps of size 256 ⍰ 256 ⍰ 64 from both H&E and IHC patches. This enabled the analysis of correlation between the two modalities in feature space through the use of learned kernels in the pretrained model. The pixel-level vector representations obtained from H&E patches depicted as *H*_*i*_ and IHC patches depicted as *I*_*i*_ 278 were utilized to train the Corr-A-Net model to learn the common representation. The individual hidden representations *h*_*H*_ = *f* (*WH*_*i*_ + *b*) and *h*_*I*_ = *f* (*VI*_*i*_ + *b*) are obtained from the multi-layer encoder for each modality for H&E and IHC respectively, where the weights of the modalities are given by *W* and *V* for H&E and IHC respectively. The multi-layer approach compared to the single-layer encoder in the original paper (36) enables to learn detailed high-level intricate features from the representations. In our work, we applied three layers of each consisting of series of convolution, batch normalization and non-linear activation using ReLU; where the filter size increased at each layer by a factor of 2. The detailed architecture of the Corr-A-Net used for correlated representation generation is depicted in Figure. 7.

**Figure 7.**
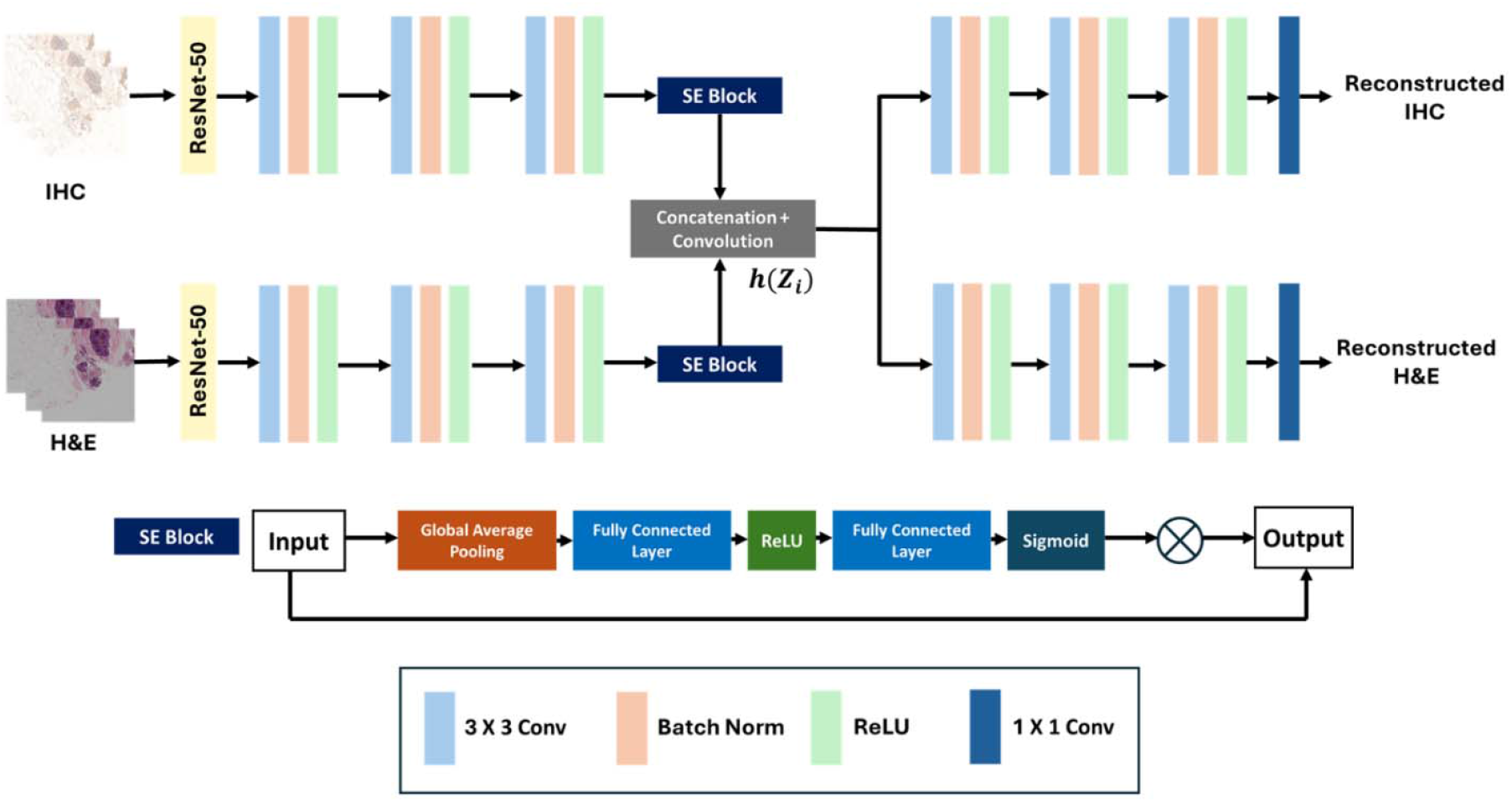
Architecture of Correlational Attention Neural Network (Corr-A-Net).

The individual hidden representations i.e. *h*_*H*_ and *h*_*I*_ are then subjected to a squeeze-excitation (SE) attention block to facilitate feature attention mechanism. The SE block aggregates the global spatial information from the modalities by applying a global average pooling operation such that each channel has an attention representation. After the squeeze step the inter-channel dependencies are learned by employing the gating mechanism through the use of fully connected layers followed by non-linear activations (ReLU) and sigmoid function. Finally, the channel-based weightage is multiplied to the original hidden representations of the modalities to get weighted attention maps. The feature fusion module takes the hidden representation from individual modalities and combine to create a shared representation which is subjected to layer of convolution and activation (depicted by *f*) in the innermost hidden layer. The shared representation is given by Eq. 1.

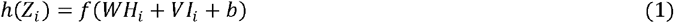

where *Z*_*i*_ = [*H*_*i*_,*I*_*i*_] denotes the concatenated vector obtained from the H&E and IHC patches. The shared representation (*h* (*Z*_*i*_)) is subjected to an identical decoder to obtain the reconstructed view.

#### Loss Function and Training of Corr-A-Net

The Corr-A-Net learns the common representations between the H&E and the IHC image patches by a joint objective function realized by maximizing the correlation between the latent representation of the two modalities (i.e. the correlation loss *L*_*Corr*_) and by minimizing the reconstruction error of the features from both modalities (*L*_*Recon*_). The reconstruction loss is further divided into two parts: self-reconstruction loss and the cross-reconstruction loss. The self-reconstruction loss is calculated by minimizing the error in reconstructing the same modality while the cross reconstruction is calculated by minimizing the error in reconstructing from cross modality. A mixture loss function composed of Structural Similarity Index Metric (SSIM) and L1-loss (Eq. 3) is used as the reconstruction objective where a hyperparameter *β* is used to control the extent between the SSIM and L1-loss (67, 68). The SSIM terms enforces perceptual and structural similarity of the reconstruction view while L1 serves as a regularizer term and helps to maintain the edges. The correlation loss is composed by the latent representations obtained from individual modalities i.e.*h*_*H*_ and *h*_*I*_ and given by Eq. 4. 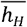 and 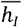 represents the mean of the latent loss representations for H&E and IHC respectively. The hyperparameter λ is used to control the extent of correlation compared to the reconstruction loss and was optimized to λ = 1.5 during the training phase.

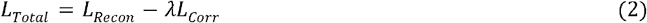

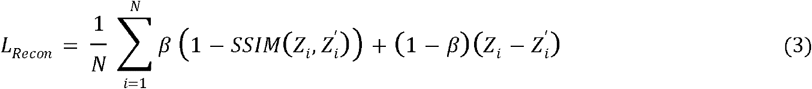

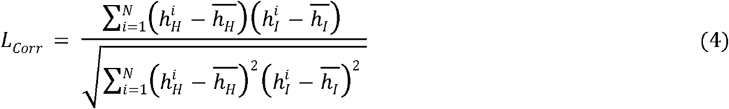

The Corr-A-Net was trained using a gradient based Adam optimizer with adaptive learning rate implemented by the Cosine Annealing algorithm. The network was trained for 250 epochs using two 48GB NVIDIA P8000 GPUs using Pytorch Lightning. After the Corr-A-Net is trained they can be used to generate latent representations without the IHC patches.

### Prediction Network and Learnable Confidence Score

We employed three deep-learning based classification networks—EfficientNet, ResNeXT-50, and Vision Transformer (ViT)—to predict HER2 status using correlated representations derived solely from H&E image patches. In addition to the prediction network, we integrated a confidence estimation network in parallel designed to learn confidence scores of the predictions. The confidence network was trained in an incentivized mechanism encouraging the network to produce reliable predictions. Furthermore, we benchmarked the performance of the correlated representation in predicting HER2 level expression to that of using IHC slides only and using both IHC and H&E combined across the channels.

#### Vision Transformer (ViT)

The ViT architecture is based on the transformer architecture which is utilized for image classification task. Unlike, traditional CNNs, ViT segments input images into a sequence of non-overlapping fixed-size patches (16 × 16 pixels) and subsequently projects these patches into learnable linear embeddings with a dimensionality of 1024. To preserve spatial information, learned positional embeddings are incorporated into the linear projections prior to their input into the transformer encoder blocks. The transformer encoder is composed of a series of twelve transformer layers, each employing multi-headed self-attention mechanisms and feed-forward neural networks (multi-layer perceptron) to process the input tokens. Layer normalization and residual connections are integrated within each layer to ensure effective gradient flow during the training process. Furthermore, dropout was applied in the transformer block to differentially switch off neurons to prevent overfitting. Finally, a fully connected layer followed by a SoftMax activation function is applied to produce probability distributions for multi-class classification tasks. This architectural design facilitates the effective modeling of long-range dependencies and complex feature representations inherent in image data.

#### EfficientNet

The EfficientNet architecture is based on the concept of compound scaling method which uniformly scales the network depth, width and resolution with set of scaling parameters. The EfficientNet utilizes a series of Mobile Inverted Bottleneck Convolution (MBConv) blocks with Squeeze-Excitation (SE) modules. In our work we utilized a variant of the EfficientNet B7 model consisting of seven blocks of MBConv ⍰ having varying degree of convolution filter size i.e. 3 ⍰ 3 and 5 ⍰ 5 filters. The MBConv block comprises an expansion phase in which the number of feature filters are increased by the expansion factor, followed by depthwise convolution where convolution is applied separately over each feature channels. The SE block then reduces the feature channels to a single value by applying global-average pulling followed by two fully connected layer with sigmoid activation to generate channel-wise weights which are multiplied to the original feature maps. Finally, the projection layer consisting of 1 ⍰ 1 pointwise convolutional blocks converts the feature maps to the same resolution as the input. The use of depthwise convolutional layers increases the time complexity of the network without affecting performance. Finally in the classification head we applied a fully connected dense layer with SoftMax activation function to obtain the multiclass probability.

#### ResNeXT

The ResNeXT architecture is based on the principle of traditional Residual Network (ResNet) model integrated with the concept of cardinality referring to the number of parallel paths within a network block. Each parallel residual sub-block consists of series of convolutions with 3 ⍰ 3 filters followed by activation, max-pooling, and feature aggregation by 1 ⍰ 1 convolution. In our model we used a cardinality factor of 32 i.e. there are 32 residual sub-blocks which are finally aggregated in the end with the input by using a long degree residual interconnection. Finally, the output is subjected to global average pooling to reduce the dimension followed by a fully connected layer with SoftMax to obtain the multiclass classification probability.

#### Confidence Estimation Network

The fundamental challenge in confidence score estimation lies in the absence of ground truth values for confidence levels. To address this, we applied an incentivized approach in which the model is rewarded to produce confidence estimates which correctly reflects the model’s ability to generate correct predictions. The confidence estimation network is coupled with the HER2 score prediction network where the penultimate layer output of the prediction network is subjected to feedforward architecture in parallel to estimate the confidence scores. The feedforward architecture consists of two fully connected layers followed by a sigmoid layer to output a confidence score between 0 and 1 (69). During training in order to give ‘hints’ to the network the SoftMax prediction probabilities obtained from the HER2 prediction network are calibrated by interpolating between original prediction and ground truth distribution given in Eq. 5.

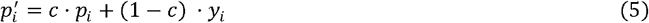

The confidence level (*c*) controls the extent of interpolation when generating the updated predictions given by *P*′.When the model is confident it outputs value such that *c* → 1, whereas if the model is unsure of the prediction, it outputs *c* → 0. We calculated our classification loss by using a weighted cross-entropy function using the updated prediction probabilities (*P*′) and ground truth targets labels (*y*). The weighted cross-entropy loss function addresses class imbalance by assigning higher weights to less frequent classes and lower weights to more prevalent classes and is depicted in Eq. 6.

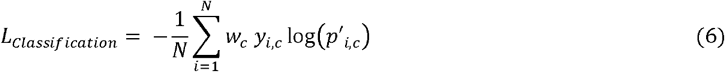

where, *W*_*c*_ is the weight vector assigned for different classes. Furthermore, a confidence loss term is applied which acts as a regularizer for the classification loss term and controls the extent of confidence level and is depicted in Eq. 7. Finally, the classification loss is combined with the confidence loss to calculate the final loss function of the network (Eq. 8). The extent of confidence loss is controlled by the hyperparameter λ.

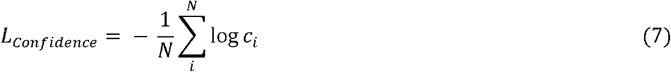

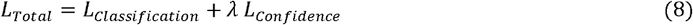

The learnable confidence score affects the dynamics of the loss function i.e. when the confidence score is high the confidence loss term becomes low, and the loss function is subjected to only weighted cross-entropy term. Whereas, when the confidence score is low the confidence loss term is high, and it penalizes the cross-entropy term.

### Performance Evaluation

The performance of the models i.e. ViT, EfficientNet and ResNeXT in predicting the HER2 level was evaluated using Weighted Precision (WP), Weighted Recall (WR), Balanced Accuracy (BA) and Matthews Correlation Coefficient (MCC) to account for the imbalanced dataset. To further assess model effectiveness, the Area Under the Receiver Operating Characteristic Curve (AUC-ROC) was calculated, and ROC curves were plotted under varying settings to illustrate the models’ discrimination capability. We also computed the Area Under the Precision-Recall Curve (PR-AUC), as this metric is robust in evaluating predictive performance on imbalanced datasets by focusing on precision-recall trade-offs and minimizing potential influence of negative class abundance.

For evaluating the model’s ability to estimate the confidence score associated with the prediction we applied the Mean Effective Confidence (MEC) metric depicted by Eq. 9.

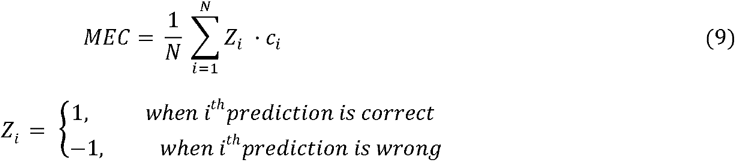

where *Z*_*i*_ denotes decision parameter whether the prediction is correct or not and *C*_*i*_ is the confidence score associated with that prediction. Thus, if the prediction is correct with high confidence or incorrect prediction with low confidence it leads to higher MEC while if the confidence value is high, but the prediction is incorrect it leads to penalization and decreases the MEC.

### Explainability Analysis

The incorporation of explainability in developing CNN models are essential as it helps in visualization of the features which are contributing to the prediction. To this end, we applied the Gradient Class Activation (Grad-CAM) method to generate representative heatmaps to interpret the model decision. We applied the Grad-CAM on the penultimate layer of the prediction network and calculated the gradients flowing backwards from the classification head to the penultimate layer. The gradients indicated the extent to which each activation in the convolutional layer influences the final classification decision. Grad-CAM essentially assigns importance weights to each feature maps based on the calculated gradients by computing the global average across the spatial dimensions. The weights assigned to each feature maps to depict the interpretability is given by Eq. 10.

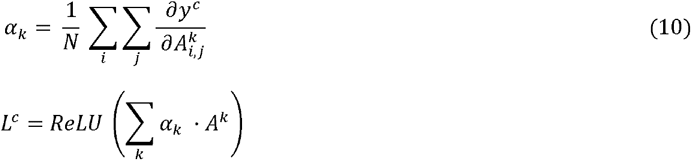

where, N is the number of pixels, *α*_*K*_ is the weight for the *K*^*th*^ feature map, y^c^ is the target class value, 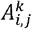 is the gradient activation at position (*i*,*j*) for *K*^*th*^ feature map. Finally, the activation maps computed for each feature is multiplied with the weights to create the Grad-CAM maps (*L*_*c*_) which signifies the relevance of each spatial location with respect to target class. The effectiveness of Grad-CAM maps lies in its ability to highlight relevant regions important for decision making. To quantitatively assess the performance of Grad-CAM maps in predicting HER2 level, we applied the Drop in Accuracy metric, which is computed as the percentage difference between the model’s prediction accuracy on the original image and the model’s accuracy when using the Grad-CAM map highlighted regions (Eq. 11). This metric indicates the relevancy of the saliency maps in prediction, with lower drops in accuracy suggesting that Grad-CAM based regions are key features driving the prediction.

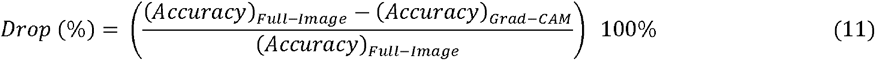

## RESOURCE AVAILABILITY

### Lead contact

Further information and requests for resources should be directed to and will be fulfilled by the lead contact, Kooresh I. Shoghi.

### Materials availability

This study did not generate new unique reagents.

### Data and code availability

- The public dataset used in this study can be downloaded from https://bupt-ai-cz.github.io/BCI/
- All original code has been deposited at Github (<u>Link</u>) and is publicly available as of the date of publication.
- Any additional information required to reanalyze the data reported in this paper is available from the lead contact upon request.

## Data Availability

All data produced in the present study are available upon reasonable request to the authors

## ACKNOWLEDGMENTS

The authors acknowledge the Center of High-Performance Computing (CHPC) at Mallinckrodt Institute of Radiology, Washington University School of Medicine for computing resources.

## FUNDING

This work was supported by NIH/NCI grants U24CA209837, U24CA253531 and internal funds provided by the Siteman Cancer Center (SCC) and Mallinckrodt Institute of Radiology (MIR). The Center of High-Performance Computing (CHPC) at Washington University in St. Louis for computing resources supported by NIH grant number S10OD025200.

## AUTHOR CONTRIBUTIONS

Conceptualization, K.D.; methodology, K.D., D.P., and S.L.; investigation, K.D., D.P., S.L., C.S., and K.I.S.; writing—original draft, K.D.; writing—review & editing, D.P., S.L., C.S., and K.I.S.; funding acquisition, K.I.S.; resources, K.I.S. and C.S.; supervision, K.I.S.

## DECLARATION OF INTERESTS

The authors have no declaration.

## Notes

### Competing Interest Statement

The authors have declared no competing interest.

### Author Declarations

The public dataset used in this study can be downloaded from https://bupt-ai-cz.github.io/BCI/

